# Provider preferences for delivery of HIV care coordination services: results from a discrete choice experiment

**DOI:** 10.1101/2021.10.21.21265350

**Authors:** Rebecca Zimba, Chunki Fong, Madellena Conte, Abigail Baim-Lance, McKaylee Robertson, Jennifer Carmona, Gina Gambone, Denis Nash, Mary Irvine

## Abstract

**Introduction:** The PROMISE study was launched in 2018 to assess and document the implementation of changes to an existing HIV Care Coordination Program (CCP) designed to address persistent disparities in care and treatment engagement among persons with HIV in New York City. We evaluated provider endorsement of features of the CCP to identify opportunities for improvement.

**Methods:** We used a discrete choice experiment (DCE) to measure provider endorsement of four CCP attributes, including: a) how CCP helps with medication adherence, b) how CCP helps with primary care appointments, c) how CCP helps with issues other than primary care, and d) where CCP visits take place (visit location). Each attribute had three to four levels. Our primary outcomes were relative importance and part-worth utilities, measures of preference for the levels of the four CCP program attributes.

**Results:** Visit location (28.6%) had the highest relative importance, followed by how staff help with ART adherence (24.3%), how staff help with issues other than primary care (24.2%), and how staff help with primary care appointments (22.9%). Within each of the above attributes, respectively, the levels with the highest part-worth utilities were home visits 60 minutes from the program or agency (19.9 utiles, 95% CI 10.7-29.0), directly observed therapy (26.1 utiles, 95% CI 19.1-33.1), help with non-HIV specialty medical care (26.5 utiles, 95% CI 21.5-31.6), and reminding clients about and accompanying them to primary care appointments (20.8 utiles, 95% CI 15.6-26.0).

**Conclusions:** Ongoing CCP refinements should account for how best to support and evaluate the intensive CCP components endorsed by providers in this study.

## Introduction

Antiretroviral therapy (ART) adherence improves clinical outcomes among persons with HIV (PWH) and reduces onward transmission of the virus [1–4]. However, in the United States, the Centers for Disease Control and Prevention (CDC) estimates that only 64.7% of persons diagnosed with HIV in 2017 and alive at the end of 2018 had achieved viral suppression (≤200 copies/mL) by the end of 2018 [5]. In New York City (NYC), 77% of PWH were virally suppressed in 2018, though stratified viral suppression rates indicate persistent disparities across multiple subgroups, including age, sex, gender, race, and transmission risk [6,7].

In 2009, the New York City Department of Health and Mental Hygiene (NYC Health Department) implemented a multi-component HIV Care Coordination Program (CCP) with the goal of improving engagement in care and treatment among the most vulnerable PWH in NYC, including those facing the additional challenges of mental health issues, food insecurity, and unstable housing [8,9]. The program has since been included in the CDC’s Compendium of Evidence-Based Interventions and Best Practices for HIV Prevention [10–12]. The initial CCP was implemented at 28 Ryan White Part A-funded agencies, reaching over 7,000 clients in less than four years. The CCP demonstrated modest benefits for viral load suppression among newly diagnosed PWH and previously diagnosed but consistently unsuppressed PWH [9,13,14]. Refinements to the CCP to enhance intervention delivery and impact were implemented in 2018, and the program will likely continue to evolve.

The PROMISE study (Program Refinements to Optimize Model Impact and Scalability Based on Evidence) was launched in 2018 to assess and document the implementation of changes to the CCP. Based on the recognition that successful program implementation depends upon both client and provider engagement, we conducted a discrete choice experiment (DCE) to understand provider preferences for specific program features. DCEs originated in econometrics [15] and increasingly are being applied to questions in the public health and healthcare settings, including HIV care and prevention [16–18]. Here we present the findings from the provider DCE.

## Methods

### Population and sampling

Ryan White Part A funding in NYC is used to fund services other than medical care, therefore our target population was comprised of non-medical providers in the core CCP positions of patient navigators/health educators, care coordinators/case managers, and program directors or other administrators at any of the 25 agencies implementing the revised CCP. All staff in those core program roles were eligible to participate. Ten agencies were community health centers, 6 were private hospitals, 3 were public hospitals, and 6 were community-based organizations. Eleven agencies were located in Brooklyn, 10 were located in Manhattan, 9 were located in the Bronx, 4 were located in Queens, and 1 was located in Staten Island. The study protocols and materials were reviewed and approved by the NYC Health Department institutional review board (IRB), which served as the IRB of record for the PROMISE study. All participants provided informed consent.

### Developing the attributes and levels

We wanted to include aspects of the CCP that could be explored in both the current provider DCE and a subsequent client DCE in order to facilitate future concordance analyses. Following from this, and in accordance with best practices for designing DCEs [19,20], we began developing a list of program features to investigate in the DCE through two client focus groups (7 participants total) and one provider focus group (5 participants) See Table, Supplemental Digital Content 1 for participant details. Both providers and clients described and expressed positivity about CCP service features, particularly those supporting non-medical issues. Providers stressed matching CCP features to each client’s particular needs.

We also considered which of the key elements of the program might be amenable to future changes, whether in program focus, intensity, or mode of delivery. The possible features and versions of those features, called attributes and attribute levels in the parlance of DCEs, were originally defined by reviewing focus group feedback and through discussion within the study team, and then refined based on feedback from PROMISE Study Advisory Board members at a meeting in June 2019. Our final design included four attributes with three to four levels each, which varied by focus, intensity, and/or mode: 1) help with adherence to ART; 2) help with primary care appointments; 3) help with issues other than primary care; and 4) where program visits happen (visit location). Black-and-white graphics (icons) were included to convey attribute levels and facilitate quick comprehension and comparison of the attribute levels across choice concepts. See Table 1 and Figure 1.

**Table 1.**
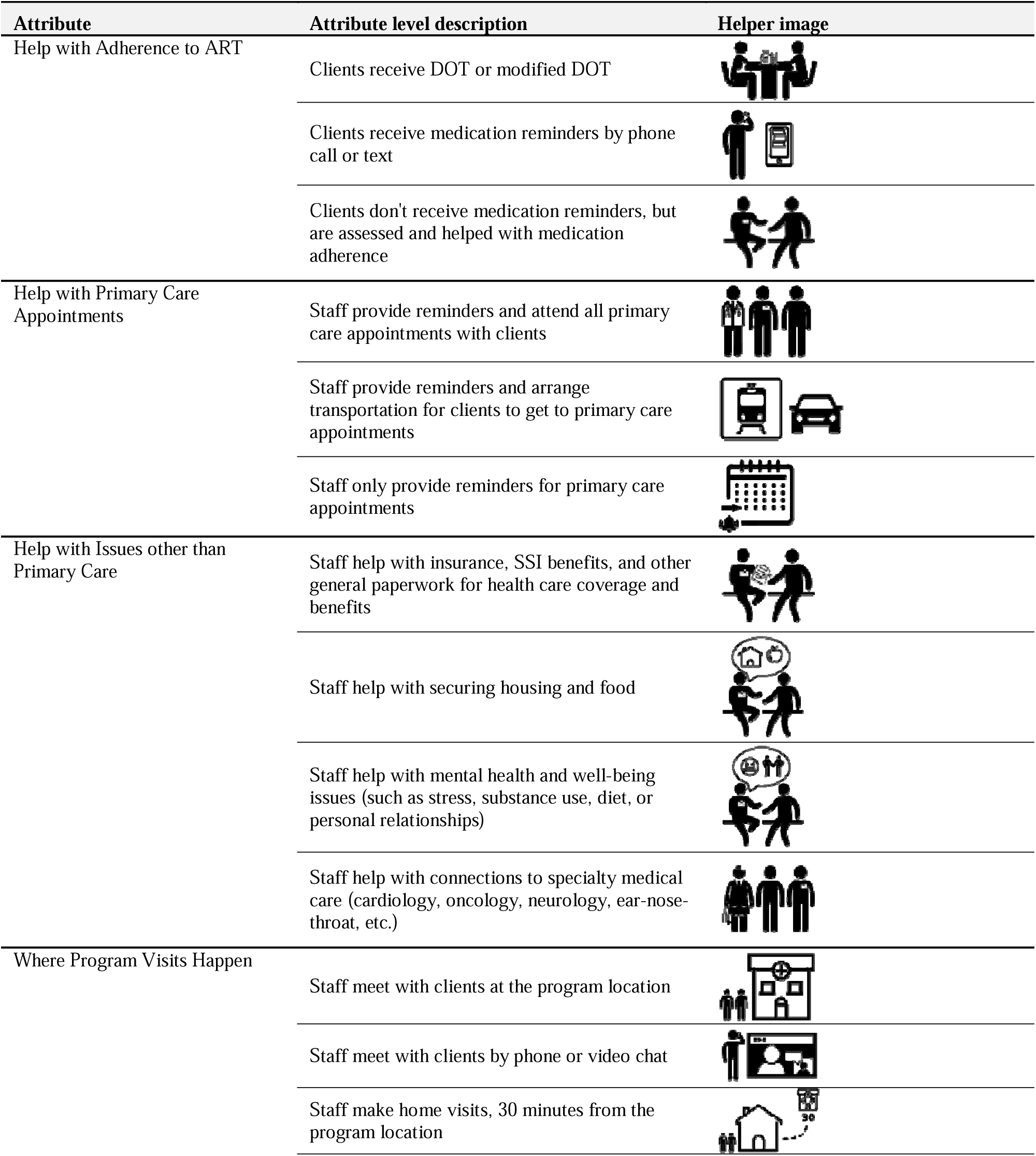
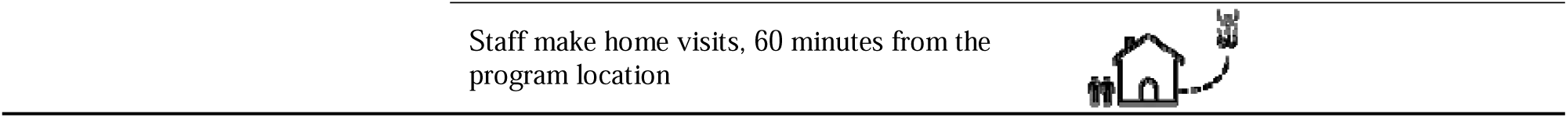
Attributes and levels of a discrete choice experiment investigating provider preferences for HIV care coordination services in New York City

**Figure 1.**
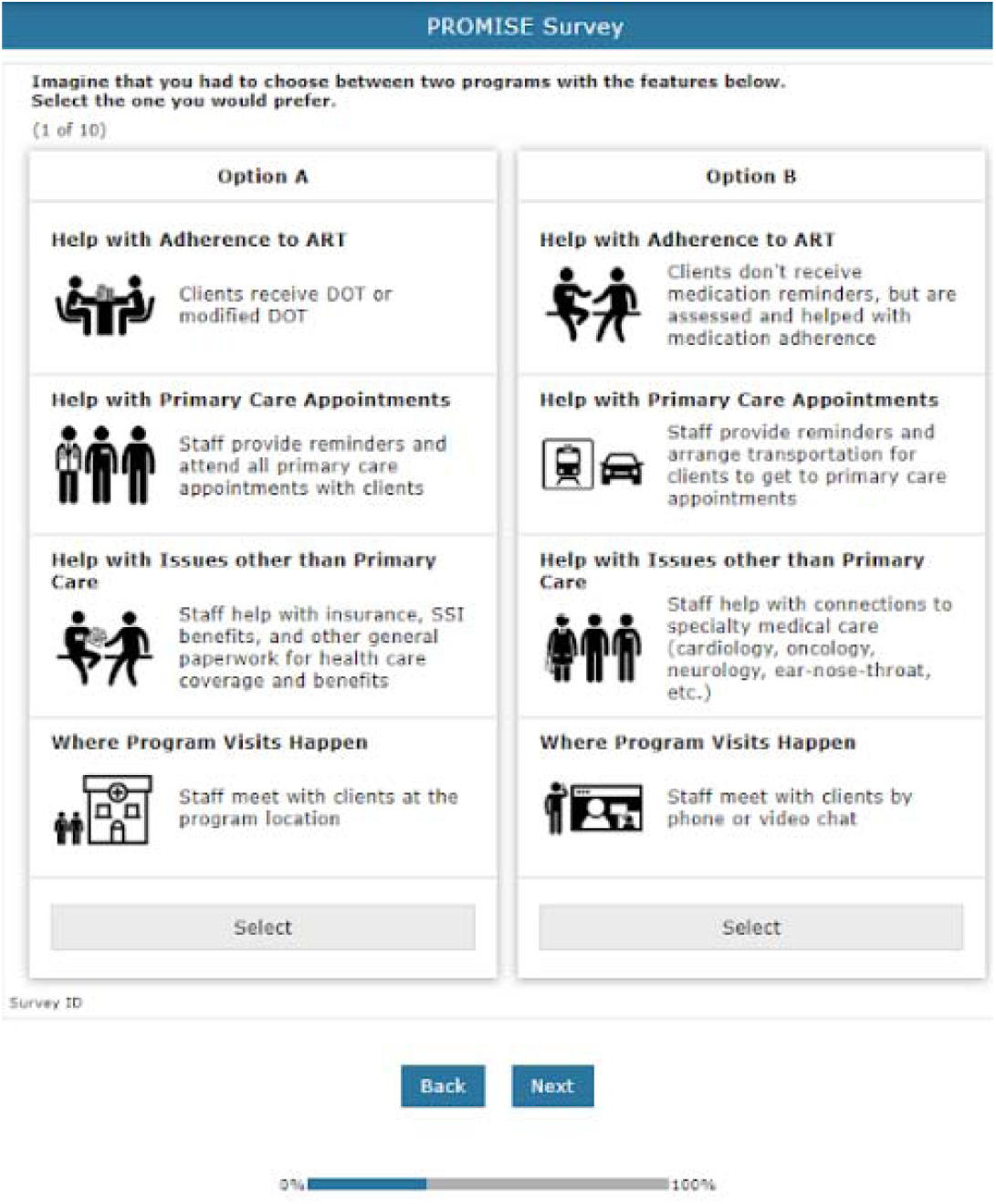
Example of discrete choice experiment (DCE) task presented to providers, desktop or laptop browser orientation.

### DCE design

The survey was designed and implemented using Lighthouse Studio Version 9.8.1 (Sawtooth Software, Provo, Utah, USA) and deployed via Sawtooth’s online survey hosting platform. The final design included ten comparison tasks, with two alternatives per choice task; to improve design efficiency and the precision of our main effects part-worth utility estimates, we chose not to include a “None” option. We used Sawtooth’s Balanced Overlap method [21,22] to generate random tasks in which each level appeared approximately the same number of times as the other levels within each attribute (level balance), some level overlap within an attribute was permitted across alternatives in the same task, and levels within one attribute were included independently of levels within other attributes (orthogonality). Our design’s relative D-efficiency was 88% compared to the D-efficiency of Sawtooth’s Completely Enumerated design, which is statistically more efficient but is less able to identify possible interaction effects due to minimal overlap between alternatives within a choice task [21]. The survey was deployed in English.

Introductory text was included to describe the attributes being investigated in the survey and explained that “Your responses will tell us what program features providers value most and what features they might like to change.” In each choice exercise, we asked providers to “Imagine that you had to choose between two programs with the features below. Select the one that you would prefer.” After the choice exercise we asked respondents about their age, race/ethnicity, and gender identity, and the length of time they had been providing CCP services.

### Sample size

The minimum sample size for estimating main effects in a DCE can be calculated as 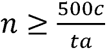, where *n* is the number of respondents, *C* is the maximum number of levels among all of the attributes, *t* is the number of choice tasks, and *a* is the number of alternatives per task [23,24]. This formula assumes each main-effect level appears at least 500 times within the survey design. The minimum sample size for our study given a maximum of 4 levels among our attributes, 10 choice tasks, and 2 alternatives per task is 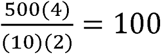, therefore, our target sample size of 150 responses was sufficient to estimate main effects.

### Data collection

In January 2020, we emailed individual survey IDs and links to the online survey to 227 providers from the 25 revised CCP implementing agencies, with a target sample of 150 completed responses. The DCE could be completed in any modern browser on a desktop or laptop computer, tablet, or phone. Participants were compensated with $25 gift cards upon completion of the survey. The survey was closed in early March 2020, before the first wave of the COVID-19 pandemic in NYC, with 152 respondents. Into the final survey data set we merged additional staff and agency descriptive data, such as staff role, agency location, and CCP budget, gathered from program liaisons and existing NYC Health Department contract records.

### Analysis

We used Sawtooth Software’s Lighthouse Studio 9.8.1 to design, administer, and analyze the survey. We estimated part-worth utilities for each attribute level using the hierarchical-Bayesian (HB) method and assuming the Random Utility Model, which posits that people choose the option that has the highest total utility for them [25]. The HB method analyzes the data at the individual level as well as the aggregate level, which yields more stable respondent-level estimates and also allows for more heterogeneity across respondents than other methods, such as traditional multinomial logistic regression [26,27].

### Part-worth utilities and relative importance

We interpreted part-worth utilities as preferences for or endorsements of particular features. We estimated part-worth utilities using effects coding, and zero-centered the estimates at the individual level in order to reduce the effect of noise on respondents’ utility ranges [15,28,29]. We calculated relative importance at the respondent level as the range in part-worth utilities for levels within an attribute over the sum of the ranges in part-worth utilities for levels in all attributes; this yielded an importance score scaled from 0 to 1 for each respondent for each attribute. We then averaged this respondent-level measure to get an aggregate-level measure of attribute relative importance. Attribute relative importance is a way to quantify the degree to which an attribute influences choices across respondents relative to the other attributes.

### Model fit and respondent data quality

We measured overall model fit using percent certainty, analogous to McFadden’s pseudo-R^2^, where values from 0.2 to 0.4 indicate a good model fit [30]. We also used the model’s overall root likelihood (RLH) to evaluate model fit. RLH is the geometric mean of the likelihood of each alternative within a choice task being selected, and ranges from 1/*n* (worst fit) to 1 (best fit), where *n* is the number of alternatives per task [31,32]. In this study, an RLH of 0.5 would indicate no model fit. We also assessed straightlining, response speed, and individual RLH values, three common indicators of respondent quality in DCEs [33].

## Results

### Respondent Demographics and Agency Characteristics

Characteristics of the 152 respondents are described in Table 2. At least one provider responded from each of the 25 CCP provider agencies (median 6 respondents, IQR 4-7 respondents). Median time to completion following consent was 7 minutes (IQR 5 minutes-12 minutes). Providers who responded to the survey were primarily Black (34%) or Latino/a (49%), identifying as women (68%), and between 30 and 49 years old (60%). Most respondents were patient navigators (65%) and had worked in Care Coordination for over two years (58%). The agencies at which most respondents worked were based in Manhattan (34%), the Bronx (28%), or Brooklyn (24%), were clinic-based (84%) (vs community-based), and had experience with the initial and revised Care Coordination model (76%).

**Table 2.**
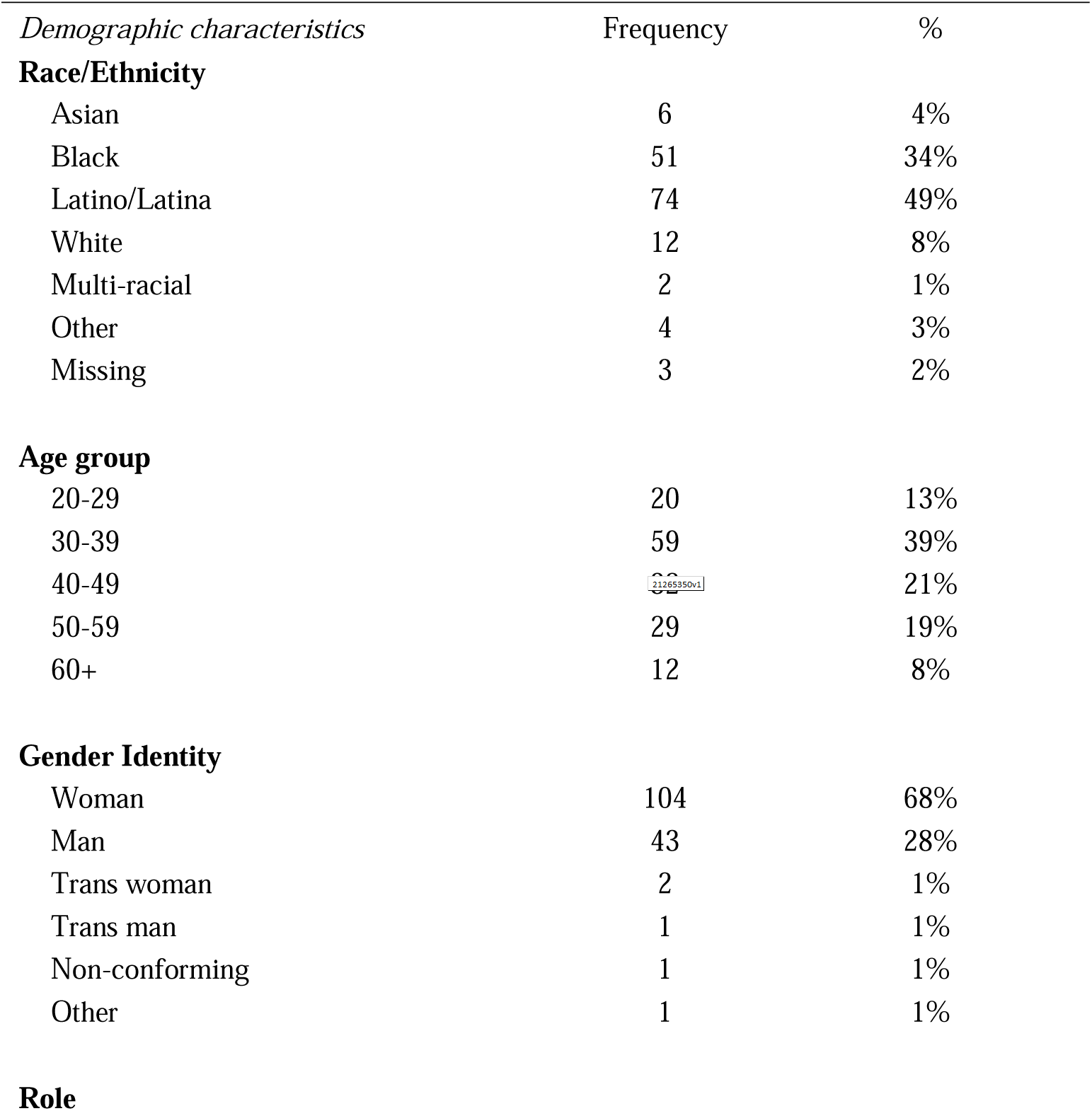
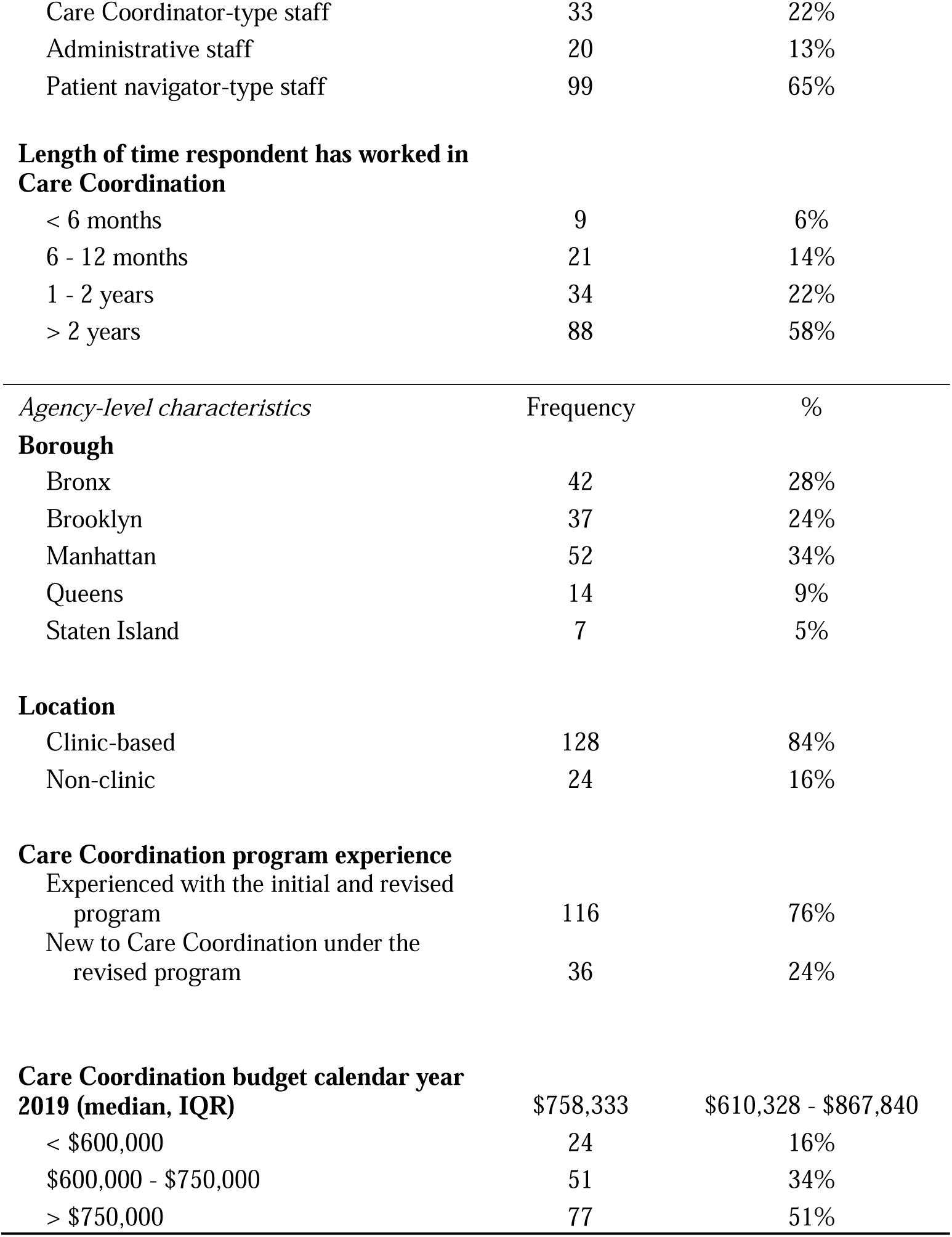
Demographic and agency-level characteristics of providers (N=152)

### Relative importance

Relative importance estimates for each attribute are shown in Table 3 and Figure 2. The attribute that had the highest relative importance was visit location (28.6%, 95% CI 27.0% to 30.3%), followed by how staff help with ART adherence (24.3%, 95% CI 22.4% to 26.1%), how staff help with issues other than primary care (24.2%, 95% CI% 22.7% to 25.7%), and lastly how staff help with primary care appointments (22.9%, 95% CI 21.7% to 24.1%).

**Table 3.**
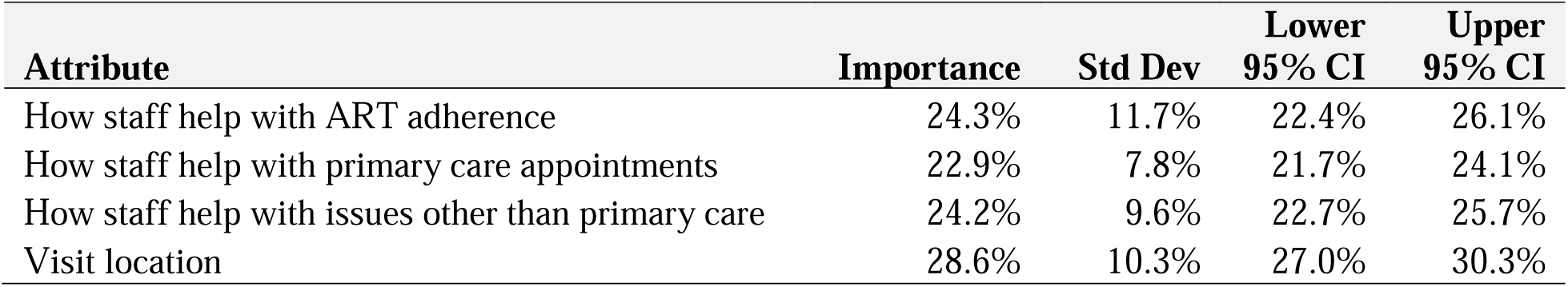
Average relative attribute importance from a discrete choice experience among providers in New York City assessing preference for HIV care coordination program features

| Attribute | Importance | Std Dev | Lower 95% CI | Upper 95% CI |
| --- | --- | --- | --- | --- |
| How staff help with ART adherence | 24.3% | 11.7% | 22.4% | 26.1% |
| How staff help with primary care appointments | 22.9% | 7.8% | 21.7% | 24.1% |
| How staff help with issues other than primary care | 24.2% | 9.6% | 22.7% | 25.7% |
| Visit location | 28.6% | 10.3% | 27.0% | 30.3% |

**Figure 2.**
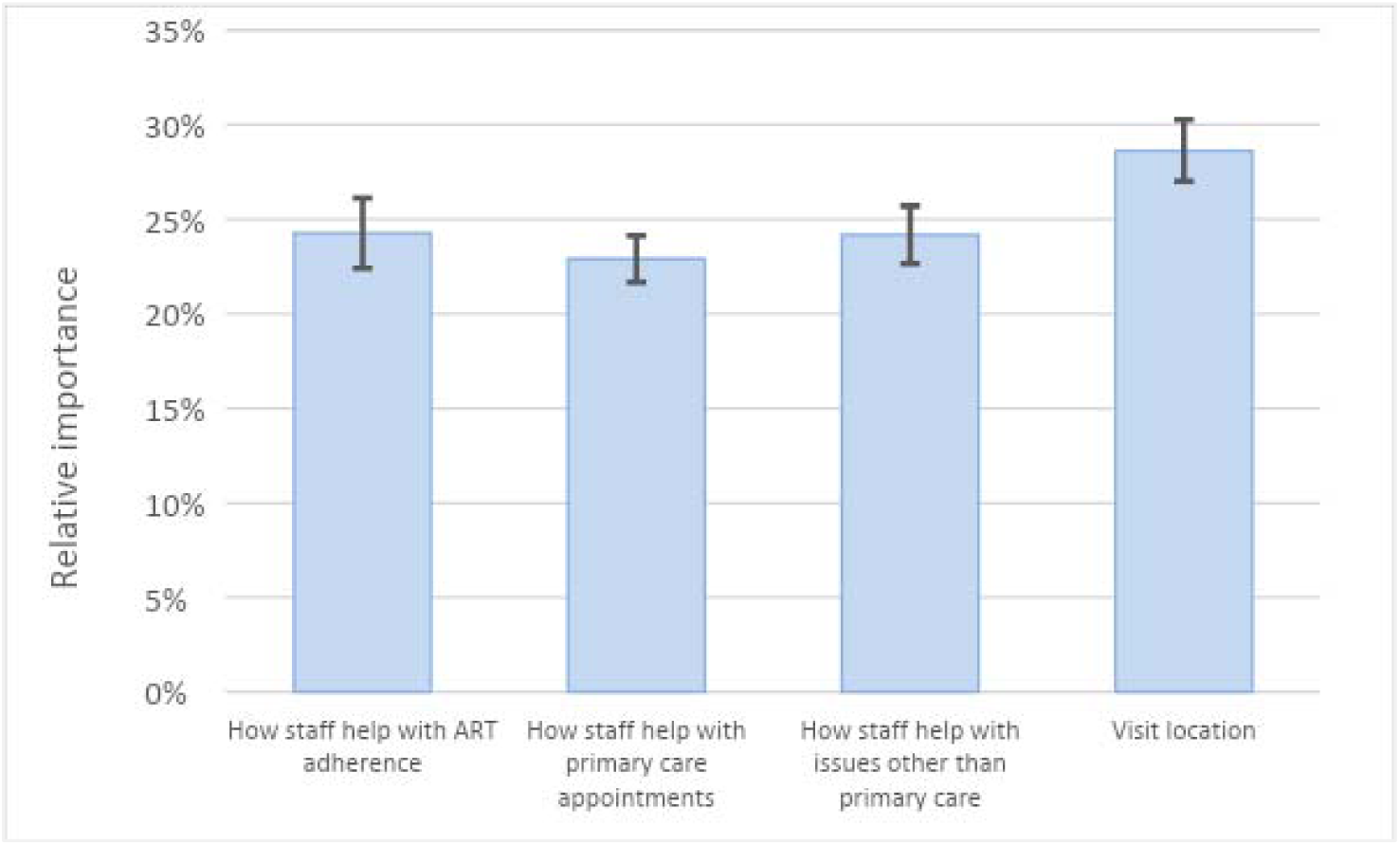
Average relative attribute importance from a discrete choice experience among providers in New York City assessing preference for HIV care coordination program features

### Part-worth utilities

Part-worth utilities of levels within each attribute are shown in Table 4 and Figure 3. The magnitude and direction of part-worth utilities indicate the strength of preference or endorsement for levels within an attribute. Providers preferred programs that included directly observed therapy as a strategy to help with ART adherence (part-worth utility 26.1, 95% CI 19.1 to 33.1), compared to reminding clients to take ART via phone or text (−5.0, 95% CI -10.2 to 0.3) and only assessing and helping with ART adherence based on responses to assessments (−21.1, 95% CI -28.5 to -13.8). Providers preferred programs that offered reminders about and accompaniment to primary care appointments (20.8, 95% CI 15.6 to 26.0) and those that reminded about and arranged transportation for primary care appointments (17.4, 95% CI 12.7 to 22.2), over those that only offered reminders about primary care appointments (−38.2, 95% CI - 43.3 to -33.0). Providers preferred programs that focused on helping clients with connections to specialty medical care for health conditions other than HIV (26.5, 95% CI 21.5 to 31.6) or with mental health and well-being (15.6, 95% CI 11.2 to 20.0), compared with programs that focused on helping with insurance, Social Security, and other benefits paperwork (2.1, 95% CI -2.6 to 6.8) or helping with securing housing and food (−44.3, 95% CI -48.9 to -39.6). Lastly, providers selected programs capable of providing home visits at locations up to 60 minutes away from the program or agency location (19.9, 95% CI 10.7 to 29.0), and home visits up to 30 minutes away from the program or agency location (8.2, 95% CI 2.6 to 13.8) more than programs in which staff only met clients at the program or agency (1.6, 95% CI -5.3 to 8.5) or met clients only via phone or video chat (−29.6, 95% CI -35.8 to -24.1).

**Table 4.** Part-worth utilities* from a discrete choice experience among providers in New York City assessing preference for HIV care coordination program features

| Attribute | Level | Utility | Std Dev | Lower 95% CI | Upper 95% CI |
| --- | --- | --- | --- | --- | --- |
| **How staff help with ART adherence** | Directly observed therapy (DOT) | 26.1 | 44.1 | 19.1 | 33.1 |
|  | Reminder via phone or text | -5.0 | 33.1 | -10.2 | 0.3 |
|  | Adherence assessment | -21.2 | 46.1 | -28.5 | -13.8 |
| **How staff help with Primary care appointments** | Remind & accompany clients | 20.8 | 32.5 | 15.6 | 26.0 |
|  | Remind & arrange transportation for clients | 17.4 | 29.9 | 12.7 | 22.2 |
|  | Remind only | -38.2 | 32.4 | -43.3 | -33.0 |
| **How staff help with issues other than primary care** | Insurance, SSI benefits & other paperwork | 2.1 | 29.3 | -2.6 | 6.8 |
|  | Securing housing & food | -44.3 | 29.3 | -48.9 | -39.6 |
|  | Mental health & well-being | 15.6 | 27.4 | 11.3 | 20.0 |
|  | Connections to specialty medical care | 26.5 | 32.1 | 21.5 | 31.6 |
| **Visit location** | At program/agency | 1.6 | 43.4 | -5.3 | 8.5 |
|  | Via phone or video chat | -29.6 | 34.8 | -35.8 | -24.1 |
|  | At clients' homes, 30 mins from program/agency | 8.2 | 35.3 | 2.6 | 13.8 |
|  | At clients' homes, 60 mins from program/agency | 19.9 | 57.5 | 10.7 | 29.0 |

\*Note: Part-worth utilities were estimated using effects coding and are zero-centered

**Figure 3.**
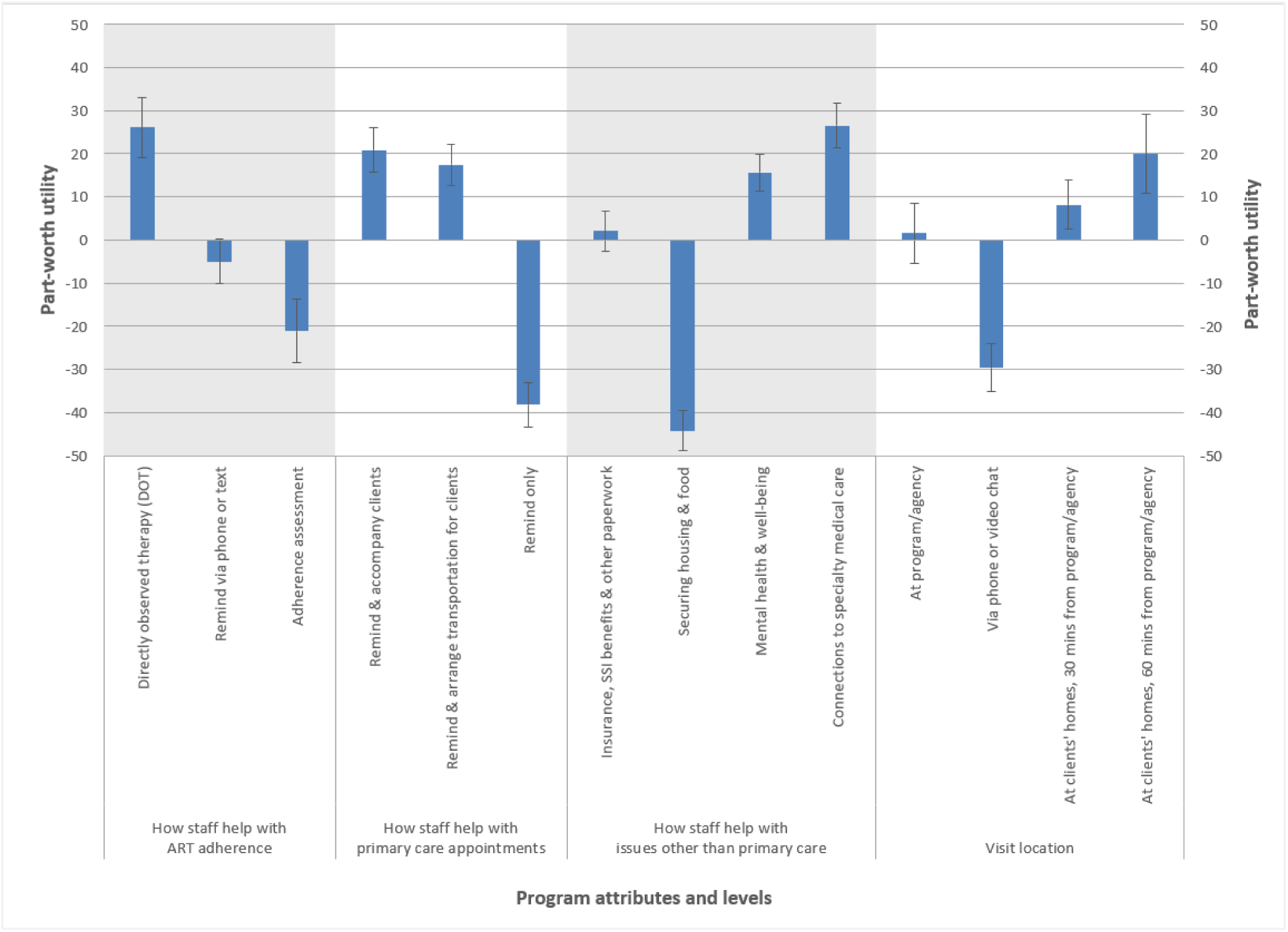
Part-worth utilities* from a discrete choice experience among providers in New York City assessing preference for HIV care coordination program features Note: *Part-worth utilities were estimated using effects coding and are zero-centered

### Model fit and respondent quality

Though we did not have any *a priori* hypotheses about interactions between attribute levels, all possible interactions were explored, and none were found to be statistically significant. Overall fit for our model measured using McFadden’s pseudo R^2^ was 0.47, and RLH for our model was 0.69, both indicating better model fit over the null model. We conducted additional analyses excluding respondents identified through straightlining, fast response time, or low individual RLH, and found that dropping potentially poor-quality respondents from the analysis made no qualitative changes to part-worth utility or relative attribute importance estimates or our interpretation of the results.

## Discussion

To our knowledge, provider preferences for features of HIV care coordination or HIV case management programs have not previously been systematically assessed. Among providers in New York City who took this survey about hypothetical variations on the HIV Care Coordination program, more intensive versions of services, such as DOT or accompanying clients to primary care appointments, were preferred over the less intensive alternatives, such as medication or appointment reminders alone.

The population for which the CCP is intended comprises PWH who have either documented risks for poor HIV outcomes, a clear history of poor HIV outcomes, or both. Our findings reflect an endorsement among participating providers of the high degree of support the program can potentially provide to these clients to help improve ART adherence. With regard to the preference for home visits, providers may have been affirming both the CCP model’s value in filling service gaps in usual agency-based care as well as an aspect of the program that makes it effective for clients, who may live up to 60 minutes from the agency location if public transportation options are limited. This may be in recognition of the value of working with clients at their homes or in the field, regardless of the distance from the clinic.

Like home-based visits, DOT is time-intensive and costly, and is uncommon in other adherence-support programs. In the original CCP, clients were assigned to particular enrollment tracks, which determined the frequency and type of services they received. After the redesign in 2018, providers had more flexibility to adjust the frequency, type, and intensity of services based on periodic assessments of individual client needs. The redesign also included the option to provide DOT virtually. This expanded access to DOT for clients and reduced barriers to providing DOT for agencies. During the 2019 grant year (March 2019 - February 2020), 14.2% of enrolled CCP clients received at least one DOT service, defined as the observation of a single dose, up from 7.7% during the 2017 grant year (March 2017 - February 2018) [34].

Coordinating specialty medical care for non-HIV health conditions or engaging clients in mental health and well-being services are activities that providers are well-positioned to undertake in the CCP, relative to other case management programs. However, our findings do not imply that providers devalue conventional case management activities. Because of how our DCE was designed, the levels within the ‘Help with Issues other than Primary Care’ attribute were mutually exclusive; in real life, a care coordination program could include support for housing and food <u>and</u> support for mental health and well-being. In fact, in recognition of the importance of supporting the whole client, the revised CCP includes reimbursable services related to helping with benefits and linking clients to housing and food services along with reimbursable services related to mental health and well-being. In this way, the CCP promotes the coordination of services across the social services and medical care systems to support the whole client. Similarly, this holistic approach to client care is reflected in the relative preferences for ways to provide more active assistance with primary care appointments.

## Limitations

Our study has several limitations which should be acknowledged. Our sample may not be representative of all Care Coordination service providers in New York City Ryan White Part A agencies with regard to provider demographics or agency characteristics, which may limit the generalizability of our findings. We were unable to compare the frequencies of characteristics between respondents and non-respondents, because individual staff demographic data are not routinely collected by the NYC Health Department. Our sample may therefore not represent the full spectrum of preferences among all Care Coordination service providers, and may suffer from selection bias. However, all 25 Care Coordination-delivering agencies and all core Care Coordination staff roles were represented among the study participants.

Our study was limited by the constraints of DCE design, which must balance obtaining valuable and actionable data with limiting respondent cognitive fatigue. While including more or other attributes and levels would have yielded different findings, the attributes and levels in our study design capture the CCP features considered important by providers and clients of Care Coordination as ascertained through our focus groups.

Finally, our ability to interpret our findings is somewhat limited by the non-specific language framing our survey and the reliance on survey self-administration. We could not tell, for example, whether some providers may have made choices based on what they thought would make the program better for clients, while others may have been thinking more about what makes the program work for themselves or their agencies. However, our larger purpose was to understand what program attributes engage providers in service delivery and identify areas where provider engagement could be improved. Since either or both of these perspectives (benefit to clients or benefit to staff and agencies) could motivate providers to deliver the services they preferred in the DCE with high fidelity, and since the most preferred services were uniformly the more intensive options presented, we believe we may interpret our findings as indicating endorsement of and positive engagement with the unique and intensive features of the Care Coordination program.

## Conclusion

Our goal in this part of the PROMISE study was to quantify providers’ preferences for features of NYC HIV Care Coordination programs and to identify discordance between the stated preferences in the study and aspects of the CCP as designed. The CCP fills gaps in an often fragmented service system through comprehensive coordination of health care and psychosocial support services. We found consistent endorsement of the intensive client-focused features that are rare in case management-type programs, such as DOT and visiting clients in their homes. We believe our findings show that providers particularly value the availability of an array of flexible Care Coordination features that have the potential to make the greatest difference for the most vulnerable clients. In response to these findings, future revisions to the CCP could aim to enhance the sustainability of the delivery of the CCP’s labor-intensive features.

## Data Availability

Data is not publicly available.

## Authors’ contributions

DN and MI conceptualized the study. ABL conducted formative work. ABL, DN, MI, and RZ collaborated on the design of the data collection tool. RZ and CF performed statistical analyses. RZ, CF and MC wrote the first draft of the paper. RZ, CF, MC, ABL, MR, JC, GG, DN, and MI contributed to interpreting the data and to the writing and revising of the manuscript.

## Additional acknowledgements

We would like to acknowledge Sarah Kulkarni for her help with logistics and securing funding for the study; Sarah Kozlowski for her help with logistics and recruitment; Kate Taylor for her contribution to the conceptualization and drafting of the DCE; and Graham Harriman and the PROMISE qualitative research team [Rachel Schenkel, Thamara Tapia, Miguel Hernandez and Honoria Guarino], for their contributions to the larger project. We would also like to acknowledge the PROMISE Study Advisory Board members for their contributions to the study (in alphabetical order by last name): Mohammed Aldhuraibi for ACACIA Network, Lori Hurley for the STAR Program at SUNY Downstate Medical Center, Tiffany Jules for Services for the UnderServed, Inc., Genesis Luciano for AIDS Center of Queens County, Cyndi Morales for the Council on Adoptable Children, and Vanessa Pizarro for COMPASS.

## Additional Files

Additional file 1: Characteristics of focus group attendees

File format: docx. Table of characteristics of focus group attendees.

